# Men’s perceptions and perceived acceptability of their female partner’s use of self-administered intravaginal therapies for treatment of cervical precancer in Kenya

**DOI:** 10.1101/2024.02.06.24302397

**Authors:** Chemtai Mungo, Konyin Adewumi, Grace Ellis, Mercy Rop, Everlyn Adoyo, Yating Zou, Lisa Rahangdale

## Abstract

**Background:** Cervical cancer continues to be a major health issue in low- and middle-income countries (LMICs). Despite increasing access to screening, access to precancer treatment remains a significant challenge in LMICs, highlighting a need for innovative, accessible, and resource-appropriate treatment approaches, including self-administered therapies.

**Methods:** A cross-sectional mixed-methods study was conducted among men aged 25-65 with a current female partner in Kisumu County, Kenya. Participants were sequentially recruited and surveyed to evaluate their understanding of HPV and cervical cancer, their views on screening and treatment, and their attitudes toward self-administered therapies. Focus group discussions with a subset of the survey participants further explored their treatment preferences and perceptions.

**Results:** Two hundred fourteen men participated in the survey, and 39 men participated in focus group discussions. The median age was 39 years, and 51% had a primary school education or less. Most (96%) were in a committed relationship, and 74% earned $10 or less daily. There was strong support for self-administered topical therapies, with 98% willing to support their partners using such treatments if available. Additionally, most participants were open to supporting necessary abstinence or condom use, though 76% believed their partners might hesitate to request condom use. When given an option, most preferred their partner to self-administer such therapies at home compared to provider administration at a health facility, citing convenience, cost-effectiveness, and privacy. Preferences varied between two potential therapies, 5-Fluorouracil (5FU) and Artesunate, based on their administration frequency, duration, and abstinence requirements. Qualitative findings largely supported the quantitative analysis.

**Conclusions:** The study demonstrates strong support for self-administered topical therapies for cervical precancer among Kenyan men. Additional research on acceptability, feasibility, and efficacy in different LMICs could pave the way for these therapies to help bridge current cervical precancer treatment gaps in these settings.

## Introduction & Background

Cervical cancer is the leading cause of morbidity and mortality among women in Sub-Saharan Africa, where insufficient healthcare infrastructure and resources have led to the profoundly disproportionate burden of a disease that is entirely preventable.^1^ Despite significant efforts to improve awareness about vaccination against human papillomavirus (HPV), which causes cervical cancer, and screening, limited access to cervical precancer treatment has remained a persistent gap in low- and middle-income countries (LMICs). In a study from Kenya, following community-based screening, only 52% of women who tested HPV-positive and were referred to a health facility for treatment ultimately received treatment within six months.^2^ Similarly, in a retrospective study on the treatment completion following cervical cancer screening among women living with human immunodeficiency virus (HIV) in South Africa, among 2072 women with abnormal pap smears between 2013 and 2018, only 174 (25.6%) underwent guideline-indicated management within 18 months.^3^ Challenges associated with precancer treatment access in LMICs include high rates of loss-to-follow-up when women screened in rural facilities are referred to urban centers where treatment is available ^4^ and high patient-to-provider ratio, resulting in significant treatment delays.^5^ Further, the risk of cervical precancer recurrence following standard surgical treatment among women living with the human immunodeficiency virus (HIV), the majority of whom reside in LMICs, remains high,^6,7^ highlighting a need to optimize treatment in this population at high risk of cervical cancer. To achieve the World Health Organization’s (WHO) target of 90% of women with cervical precancer receiving adequate treatment globally by 2030,^8^ there is an urgent need for innovative, feasible, and cost-effective strategies to close the precancer treatment gap for women in LMICs.

The use of topical, self-administered therapies for cervical precancer treatment offers a feasible and cost-effective opportunity to bridge the current treatment gaps in low-resource settings. While no non-surgical therapies are currently approved for the treatment of cervical precancer, the feasibility, acceptability, and efficacy of topical therapies have been demonstrated in several studies in high-income countries,^9–12^ including randomized trials.^13–16^ Two of these medications, 5-Fluorouracil (5FU) and Artesunate, are on the WHO’s List of Essential Medications,^17^ are generically available in LMICs, and could be repurposed as self-administered precancer therapies in LMICs if supported by local evidence. Topical therapies for treating cervical precancer, many of which can be self-administered intravaginally, offer a solution to overcome the difficulties faced with facility-based treatments in LMICs. These challenges include limited access to skilled providers, often located in referral facilities that are far from rural areas where most women reside.^5^ Our team has demonstrated high acceptability of intravaginal topical therapy among a cohort of women undergoing cervical cancer screening, with a preference for self-application of treatment at home compared to provider-application of treatment at a clinic.^18^ Despite these data, there are no studies evaluating men’s perceptions of their female partner’s use of self-administered topical therapies for cervical precancer treatment in sub-Saharan Africa. Male partner support has been shown to improve reproductive health outcomes among women in LMICs. In contrast, a lack of male partner support is a well-documented barrier to the uptake of cervical cancer prevention-related interventions in sub-Saharan Africa.^19–24^ Specific to the potential use of intravaginal therapies for cervical precancer treatment, male partner support is crucial, as these therapies often require periods of abstinence during their use. Though there is limited literature on intravaginal topical therapies for the prevention of cervical cancer in LMICs, studies on the use of intravaginal microbicides for HIV prevention among women in Sub-Saharan Africa have shown that male partner support is essential to uptake and adherence.^25–27^ To inform feasibility studies in LMICs, we carried out a mixed-methods study in Kenya among men with active female partners to understand their knowledge of HPV and cervical cancer screening, their perspectives, and their perceived acceptability of topical, self-administered therapies for cervical precancer treatment.

## Methods

### Study design, recruitment, and setting

We conducted a cross-sectional, mixed-methods study in Kisumu County, Kenya, between March and December 2023. Eligible participants were men ages 25 – 65 years with a current female partner. Participants were recruited primarily from outpatient HIV clinics in Kisumu County, Kenya, as well as from women undergoing cervical cancer screening or precancer treatment who could invite their male partners to participate in the study. A convenience sampling technique was utilized, where eligible participants were invited to participate and sequentially enrolled during the study period.

Kisumu County is one of 47 administrative units in Kenya,^28^ a country of 55.1 million in East Africa.^29^ Kisumu County is among the highest HIV burden regions in Kenya, with a 17.5% prevalence rate, compared to a national average prevalence of 4.9% in 2018.^30^ In Kenya, cervical cancer is the leading cause of cancer death for women, with an estimated 3,200 deaths in 2020.^31^ In Kenya, the National Cancer Guidelines recommend cervical cancer screening for women ages 25 – 65 years. Consistent with the WHO guidelines,^32^ treatments for cervical precancer in Kenya are offered via provider-administered ablation or excisional methods.

### Quantitative data collection and analysis

All participants completed an interviewer-administered closed-ended questionnaire that evaluated participant knowledge of cervical cancer risk factors, screening, and treatment. Participants were also asked close-ended questions to assess their perceived acceptability of their female partner’s use of topical therapies for cervical precancer treatment, which can be self-or provider-administered. Sociodemographic characteristics were also included in the questionnaires. The questionnaires, which included sociodemographic characteristics, were administered by trained research assistants in a private room and in the participants’ preferred language, either English, *Dholuo*, or *Swahili*. Quantitative data was entered directly into REDCap’s secure survey platform for analysis. Descriptive statistics were used to summarize questionnaire data. All statistical analyses were performed using R version 4.1.0 (Vienna, Austria).

### Qualitative data collection, transcription, and translation

A subset of the survey participants were invited to participate in focus group discussions (FGDs), which were scheduled at a later date. All participants who completed the survey could participate in the FGDs. However, a preference was given to men whose female partners had a history of cervical precancer treatment, based on self-report. Thirty-nine men participated in five FGDs, with recruitment into the FGDs occurring sequentially. A sample size of five focus groups was determined a priori based on evidence suggesting most themes are captured in three to six focus groups.^33^ In the FGDs, we adopted a constructivist paradigm to understand men’s views regarding cervical cancer screening prevention, treatment of HPV and cervical precancer, and their perceived acceptability and support of their female partners’ use of self-administered topical therapies for cervical precancer treatment were it to be recommended by a health provider. Constructivism posits that understanding is derived (i.e., constructed) based on one’s perceptions, experiences, and social contexts.^34^ Therefore, we hypothesized that men’s acceptability of topical, self-administered therapies is based, in part, on their experiences (such as having a female partner who had ever been diagnosed with HPV or cervical precancer or cancer and prior experiences with the health system) and their social contexts (such as relationships with sexual partners).

The FGDs included five to eight participants each and were held in facilities that were geographically convenient to the recruiting clinics. The FGDs were conducted in the two most spoken local languages (*Swahili* and *Dholuo*) and were guided by several domains of inquiry: 1) baseline knowledge of HPV and cervical cancer screening and prevention; 2) perception of the female partner’s risk of HPV or cervical cancer; 3) prior experience of a female partner undergoing cervical precancer treatment; 4) perceived support for and acceptability of female partners using self-administered topical therapies for HPV or cervical precancer treatment; and 5) perceived barriers and facilitators of the use of topical therapies among female partners. During the FGDs, participants were introduced to the two potential topical therapies for precancer treatment for which data are available 5-FU ^14^ and Artesunate.^10^ Information regarding the two therapies was provided to participants, including details on their potential frequency of use (5-FU once every other week for eight applications, Artesunate daily for five days for three cycles), abstinence requirements (two to three days of abstinence after each 5-FU application and none for Artesunate), and the recommendation of consistent contraception use while using both therapies. The participants’ perceptions and perceived support of their female partner’s use of topical therapies were explored in a hypothetical scenario in which the participants’ female partners needed precancer treatment and a topical therapy was recommended, with discussions about male partner support of the various requirements, including abstinence or contraception requirements. Each FGD lasted approximately 90 minutes. To promote anonymity, participants identified themselves by respondent number (e.g., R1, R2…R8). All FGDs were audio recorded, transcribed verbatim, and translated from *Swahili* and *Dholuo* to *English* by the FGD moderators. The two moderators cross-checked the translations to confirm that they captured all discussions that were recorded.^35^

For qualitative data analysis, a codebook was created during the coding process through agreement among two coders (GZ, SKG) who read and coded two of the five FGD transcripts to gain a sense of the topics covered and group discussions. All FGD transcripts were coded using the developed codebook. To ensure inter-coder agreement, a subset of transcripts was randomly selected, and codes were compared for agreement; discrepancies were resolved through discussion and consensus, with revisions documented in the codebook. Content analysis was performed using NVivo V1.71. Because the proposed topical treatment is novel in this context, our analysis involved using qualitative description, which is well-suited for increasing understanding in an area with limited knowledge.^36^ As this approach stays ‘close’ to the data with minimal interpretation, qualitative description supported our intent of straightforward description of participant experiences that included describing and relaying perspectives using participants’ own experiences and language. Coding reports were generated from NVivo and carefully reviewed to identify themes relating to male involvement and support of cervical precancer treatment. In this manuscript, we focus on men’s preferences of potential topical therapies based on their characteristics, as discussed in the FGDs.

### Research team

The research team included the principal investigator (CM), a Kenyan-born practicing obstetrician/gynecologist with seven years of experience, graduate students in medicine, social work, and public health (KA, SKG, GZ), a senior qualitative investigator with nearly 20 years of experience in qualitative methods and health services research (RMF), and a senior gynecologist with over 15 years of experience studying topical therapies in the US context (LR). The focus groups were moderated and transcribed by two qualitative research assistants from the local community who spoke local languages and were conversant with the local culture (EA, JO). The moderators had training in qualitative research, prior experience conducting FDGs, familiarity with the local context, and fluency in the local languages. The moderators also received additional training from the principal investigator on the study topic, protocol, and informed consent.

### Ethical Considerations

The study was approved by the ethics review boards at Maseno University School of Medicine in Kenya and the University of North Carolina at Chapel Hill in the US. All participants provided consent prior to participation in the study.

## Results

### Demographics

A total of 214 surveys were administered among men with current female partners in Kisumu County, Kenya. The median age of respondents was 39 years old (Table 1). Fifteen percent of study participants had less than a primary school education; 36% had completed primary school; 26% had completed secondary education; and 23% had a college education or higher. The majority of respondents (56%) were self-employed in the informal sector, 37% had formal employment, and 15% were unemployed. The majority, 74%, had a daily income of less than $10, and 96% identified with the Christian religion. The majority sought health information from health facilities (69%), followed by the radio (17%). Nearly all were married to or living with their partner (96%), and most had fathered three or more children (64%). The majority (73%) self-reported as positive for HIV, and 39% reported a history of sexually transmitted infection. Thirty-nine men participated in five focus group discussions (FGDs). The mean age of participants was 42.5 years (interquartile range: 37.5 years to 48 years) (data not shown). Most FDG participants, 31 (79.5%), had a female partner with a history of cervical precancer treatment.

**Table 1.**
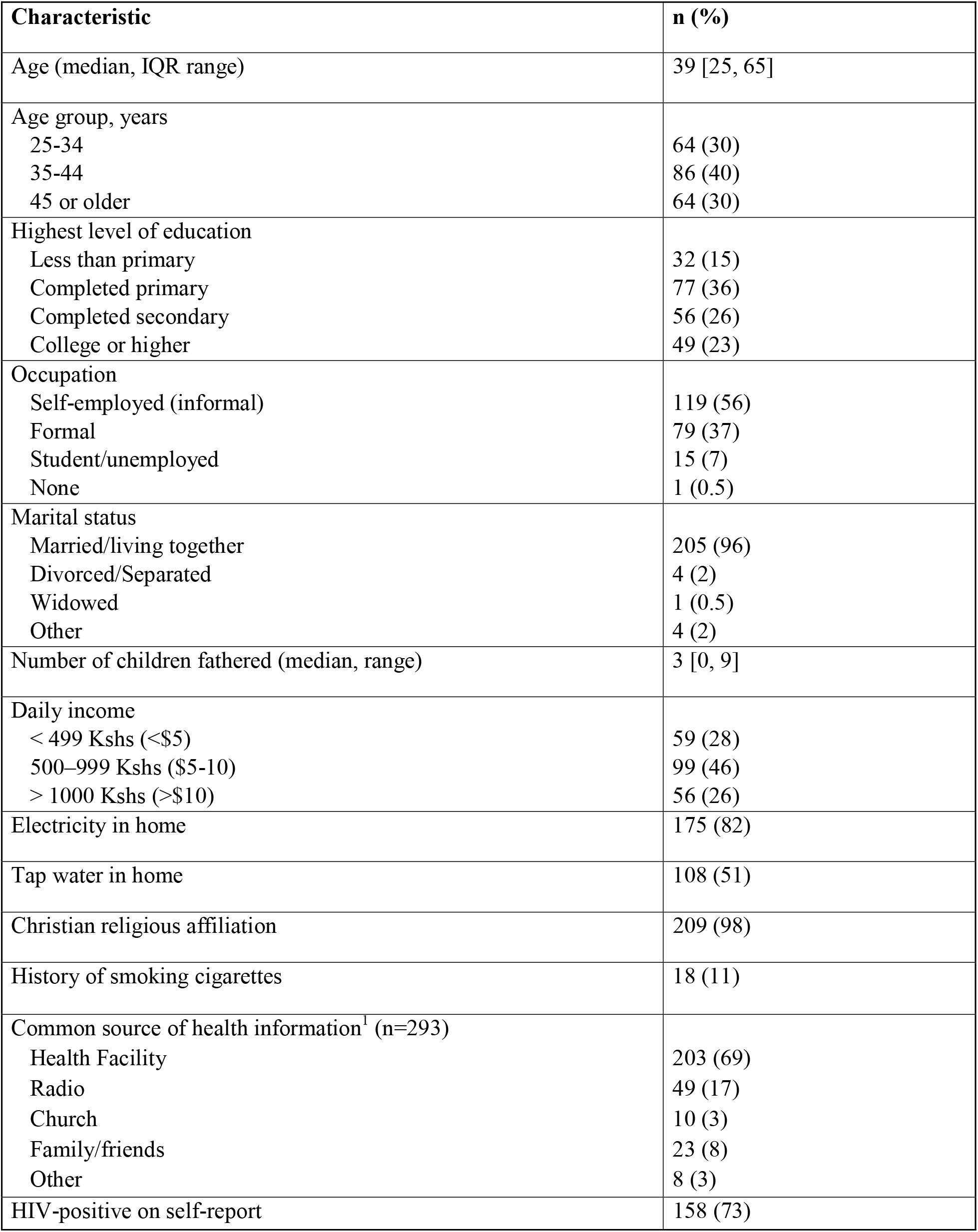
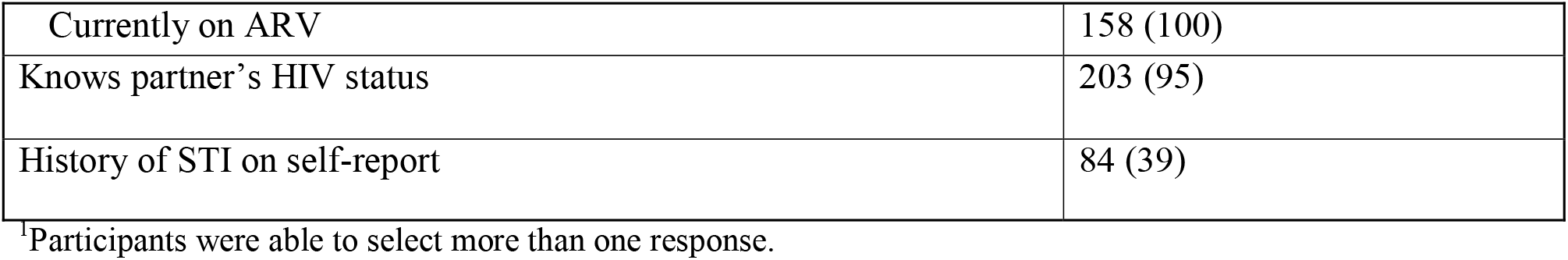
Sociodemographic and clinical characteristics of partnered men ages 25 – 65 in Kisumu County, Kenya, n=214.

| Characteristic | n (%) |
| --- | --- |
| Age (median, IQR range) | 39 [25, 65] |
| Age group, years |  |
| 25-34 | 64 (30) |
| 35-44 | 86 (40) |
| 45 or older | 64 (30) |
| Highest level of education |  |
| Less than primary | 32 (15) |
| Completed primary | 77 (36) |
| Completed secondary | 56 (26) |
| College or higher | 49 (23) |
| Occupation |  |
| Self-employed (informal) | 119 (56) |
| Formal | 79 (37) |
| Student/unemployed | 15 (7) |
| None | 1 (0.5) |
| Marital status |  |
| Married/living together | 205 (96) |
| Divorced/Separated | 4 (2) |
| Widowed | 1 (0.5) |
| Other | 4 (2) |
| Number of children fathered (median, range) | 3 [0, 9] |
| Daily income |  |
| < 499 Kshs (<\$5) | 59 (28) |
| 500–999 Kshs (\$5-10) | 99 (46) |
| > 1000 Kshs (>\$10) | 56 (26) |
| Electricity in home | 175 (82) |
| Tap water in home | 108 (51) |
| Christian religious affiliation | 209 (98) |
| History of smoking cigarettes | 18 (11) |
| Common source of health information<sup>1</sup> (n=293) |  |
| Health Facility | 203 (69) |
| Radio | 49 (17) |
| Church | 10 (3) |
| Family/friends | 23 (8) |
| Other | 8 (3) |
| HIV-positive on self-report | 158 (73) |
| Currently on ARV | 158 (100) |
| Knows partner's HIV status | 203 (95) |
| History of STI on self-report | 84 (39) |

<sup>1</sup>Participants were able to select more than one response.

### Knowledge of HPV and cervical cancer

We evaluated survey participants’ knowledge of HPV and cervical cancer and their attitudes towards their female partners undergoing cervical cancer screening or precancer treatment (Supplemental Table 1). The vast majority (98%) of survey participants had previously heard of cervical cancer and were aware that women can undergo screening for it. Most respondents did not know anyone with cervical precancer or cancer (66%), but many (65%) felt their partner may be at risk for the disease, and 84% reported their partner had undergone screening at least once. Among the 38% of respondents who had heard of HPV previously, 85% knew HPV could be transmitted through sexual intercourse, 63% knew that both men and women could be infected with HPV, 96% knew HPV causes cervical cancer, and 90% knew that having HIV increased a woman’s risk of getting HPV or cervical cancer. However, only 61% of respondents were aware that cervical cancer could be prevented through vaccination or screening. Almost all participants responded that they support cervical cancer screening for their female partners (99%) and would support them if they needed treatment (99%). Only 5 (2%) felt that a cervical cancer diagnosis would negatively affect their relationship.

### Men’s perceived support of topical therapies

Survey respondents were asked about their perceived acceptability of their female partner’s use of topical therapies for cervical precancer treatment and whether they were available and recommended to them. Nearly all (98%) reported they would support the use of self-administered intravaginal treatment at home by their partner (Table 2). Similarly, the vast majority would be willing to abstain from sex (99%) or use condoms (98%) for a certain period during their partner’s treatment. A majority (76%) agreed that women undergoing self-administered treatments may be afraid to ask men to use condoms due to fear of violence. However, only 10% of respondents felt their own partner would be uncomfortable asking them about the need to abstain while she was using a topical treatment. Most respondents (79%) did not know what a tampon was at the time of the survey. However, after the research assistant provided a brief explanation and showed them a picture of a tampon, 97% stated that they would be comfortable with their female partner using a tampon as part of topical therapy treatment.

**Table 2.** Self-reported acceptability of female partner’s use of topical therapy for cervical precancer treatment among partnered men in Kisumu County, Kenya, n=214.

| Question | Yes (%) | No (%) | Unsure (%) |
| --- | --- | --- | --- |
| Would support use self-administered intravaginal treatment at home | 210 (98) | 2 (1) | 2 (1) |
| Willing to abstain from sex for a certain period during treatment | 211 (99) | 1 (0.5) | 2 (1) |
| Willing to use condoms for a certain period during treatment | 209 (98) | 3 (1) | 2 (1) |
| Partner would be comfortable asking for condom use because of treatment | 202 (94) | 4 (2) | 7 (3) |
| Would support use of dual contraception to avoid pregnancy during treatment | 211 (99) | 1 (0.5) | 2 (1) |
| Women may be afraid to ask for condom use due to violence | 163 (76) | 23 (20) | 8 (4) |
| Possible that partner may feel uncomfortable asking for abstinence | 21 (10) | 186 (87) | 7 (3) |
| Knows what a tampon is | 41 (19) | 170 (79) | 3 (1) |
| Would be comfortable with partner using tampon as part of treatment | 207 (97) | 3 (1) | 4 (2) |

### Men’s preference regarding the location of administration of topical therapies

Survey participants were asked about their location preference for their female partner’s use of intravaginal topical therapy, with options of self-administration at home or provider-administration in a health facility. Forty-nine percent of participants preferred self-administration at home, 32% preferred provider-administration in a health facility, and 19% were unsure or did not have a preference (Table 3). Of those that preferred self-administration at home, 50% chose this option as it was cheaper (less money spent on transportation to clinics), 40% believed this was easier compared to provider-administration, and 10% had other reasons. Among respondents who preferred provider-administration, 56% felt unsure that their partner could correctly self-apply the medication, 35% believed provider-administration would be safer than self-administration, and 8% had unknown reasons for their preference.

**Table 3.** Preferences for topical therapy for cervical precancer treatment among men with female partners in Kisumu County, Kenya, n=214.

| Characteristic | n (%) |
| --- | --- |
| **Preference for location of topical therapy application** |  |
| Self-administration at home | 105 (49) |
| Provider-administration at clinic | 69 (32) |
| No preference or unsure | 40 (19) |
| Reason for self-administration at home preference<sup>1</sup> (n=135) |  |
| Easier to administer | 54 (40) |
| Cheaper/less transport | 67 (50) |
| Other | 14 (10) |
| Reason for preference for provider-administration in clinic<sup>1</sup> (n=85) |  |
| Safer | 30 (35) |
| Unsure if partner can safely-apply correctly | 48 (56) |
| Other | 7 (8) |
| **Preference for type of topical therapy based on two provided options<sup>2</sup>** |  |
| Based on frequency of dosing and treatment duration |  |
| Preference for 5FU | 123 (57) |
| Preference for Artesunate | 91 (43) |
| Based on abstinence and contraception requirements |  |
| Preference for 5FU (Option 1) | 119 (56) |
| Preference for Artesunate (Option 2) | 95 (44) |
| Preference for treatment duration with every other week application |  |
| 8-week treatment course with 4 doses | 194 (91) |
| 8-week treatment course with 4 doses | 14 (6) |
| No preference | 6 (3) |

<sup>1</sup>Participants were able to select more than one response.
<sup>2</sup>Option of self-administered 5FU vs Artesunate presented as potential options. 5FU described as use every other week for 8 applications (16 weeks total), requiring abstinence for 2-day after use. Artesunate described as daily use for 5 days straight, followed by a week off (Cycle 1), repeated for 3 cycles, with no abstinence requirements.

During the FGDs, when presented with the option of self-administered topical therapy at home versus provider-administered services at a clinic, all 39 FGD participants preferred self-administration at home application. Reasons mentioned for preferring self-administered therapy at home included distance to the clinic, cost of transportation, clinic wait times, and privacy.

> *I think this treatment for cream is good, so long as women are trained on how to use it, it will be easy because transport is not easy, because it can reach a time when one is busy with other things, even travelling to the hospital becomes a burden, so if she is given this cream to have in the house, it will be much easier. – (R3, FGD3) I would prefer cream because going to the hospital will take time and fare but the cream you apply in your house at your own time. – (R5, FGD3)*

> *The cream for use at home is better because going to the hospital is not easy, sometimes when you go to the hospital there is a long queue before seeing the doctor and maybe you have something to chase back, this can inconvenience someone. So, the one used at home is good. – (R4, FGD 3)*

Of the FGD participants who cited *privacy* as their reason for preferring self-administered at home, the discreet nature of self-administration was often emphasized, as well as the perception that self-administration could address cultural barriers related to provider-administered therapies:

> *Kenyan women will use it because it’s easy to use, you have it in the house and you just apply, its good because it’s a secret between you and your partner so my request is that the government should avail it in the local chemists. – (R7, FGD3)*

> *Many women will like this cream because of privacy, many would not like their private parts to be touched and this cream would work for them. – (R3, FDG3)*

> *Cream is good because the other three methods [cryotherapy, thermal ablation, loop excision, some women does not like someone to touch their private part but with the cream, you just use it on your own in the house in the house. -(R4, FGD 3)*

While no FGD participant explicitly mentioned a preference for provider-administration in a facility, several participants emphasized the need for sufficient education from a healthcare provider about proper self-administration of such therapies or an initial period of provider-administration as part of the education process.

> *I would advise women or my wife to go the doctor for explanation on how to use the cream so that she will be applying it by herself at home. This would save her even the time for going to the hospital from time to time. When the doctor has showed her how to do it, this will be very good. – (R4, FGD5)*

> *I think when she comes to the clinic, the first days of treatment, she should be orientated to the cream and how to apply to by herself, because the doctor will apply for her the first early days of treatment. Therefore, I prefer that from the beginning, the doctor applies cream for her but some time and orientation, she will be applying it all by herself. – (R3, FGD5)*

### Men’s preferences for topical therapy: 5FU and Artesunate

During the survey, 5FU and Artesunate were presented as potential options for self-administered intravaginal therapies that may be available to women, with the characteristics of each therapy described using visual aids, including frequency of use and abstinence requirements. Participants were then asked which of the two therapies they would prefer for their female partner to use, considering the distinct characteristics of each. Specifically, they were queried on their preferences in terms of the frequency of use and treatment duration of each, as well as their abstinence and condom use requirements. 5FU is administered biweekly for a total of 8 applications over 16 weeks, and women must abstain from sex for two days following each application. In contrast, Artesunate requires daily administration for five days, followed by a drug-free week, repeated for three cycles, and abstinence following use is not required. When surveyed on their preferred choice between the two therapies, considering frequency and duration of use, 57% of participants favored 5FU (Table 3). When considering the requirements for abstinence and contraception, 56% of participants expressed a preference for 5FU. This preference was despite the need for 48 hours of abstinence after each application of 5FU, compared to Artesunate, which does not require abstinence for a set period after use. However, like with 5FU, condom use is recommended during the course of Artesunate treatment. Only 44% preferred Artesunate under these considerations. Specific to 5FU, participants were asked whether they preferred a shorter treatment duration compared to a longer treatment duration if they are both equally effective. The vast majority, 91%, noted a preference for shorter treatment duration (an 8-week treatment course with four applications over a 16-week treatment course with eight applications).

During the focus groups, reasons given for why participants preferred 5FU were related to men’s perceptions that it would be easier for women to remember to use a therapy once every two weeks compared to daily.

> *I feel that since the woman has other activities in addition to taking drugs, you know women have women groups for merry-go-round, and so if you are to put her on full week medication, it won’t be possible and therefore the one for once a week would work well for her. -(R7, FGD5)*

Another FGD participant thought that longer duration may be related to efficacy, hence favoring 5FU, stating that the shorter duration of Artesunate therapy may be a better fit for those who cannot abstain from sex for long.

> *I feel the women should go for one that runs for sixteen weeks, my reasoning is that the one for five [days] will not work as effectively as the one for longer period. My thinking is that the one for short period is for guys who cannot abstain from sex for long. But the best way in my opinion is that we should have ample time for healing and therefore the choice of sixteen weeks. Otherwise, if you cannot abstain for a long period of time, then you better take your wife to stay in rural home during healing time and you remain in town working for income. – (R5, FGD1)*

Reasons mentioned for preferring Artesunate to 5FU in the FGDs included not needing to maintain abstinence for several days following Artesunate use and the shorter duration of treatment during which condom use is recommended.

> *I prefer the one for five days and get over the treatment within [six] weeks unlike the other one which staggers for sixteen weeks. Sixteen weeks is such a long time, it is about four months and somebody is bound to forget that the wife is on treatment and you won’t use condom. Again, it is not normal to be using condom with your wife every time you want to have sex. So, I feel the one for five days then a one-week break is better. – (R3, FGD1)*

> *Like any other man or one of the men, I would choose number two called Artesunate because you can have sex on day of treatment before you apply, this cannot bring issues in the house because after sex then you can explain to the husband that you will now start using the drug. – (R2, FGD3)*

Another FGD participant believed that Artesunate’s daily dosing schedule would be easier for his wife to remember, potentially leading to better adherence.

> *There is great compliance to drug taking because she will be aware that for this week she is on cream and so even her work schedule will be aligned to [the] cream application and [she will] create time for it. She won’t forget and she will very much comply. Since it gives room for resting for a week, this will be good and even on my side as her husband, I will know all her actions and comply with all the treatment requirements. – (R4, FGD5)*

## Discussion

In a mixed-methods study on men’s attitudes toward their female partners’ use of topical therapies for cervical precancers in western Kenya, we find overwhelming support for these treatments. Among 214 surveyed men, most of whom had heard of cervical cancer before, nearly all stated they would support their partner’s use of topical therapy if it was recommended. Despite most having limited education and low income, many were willing to practice sexual abstinence and use condoms during their partner’s treatment. However, concerns were raised about women’s reluctance to request condom use due to possible fear of violence. About half of the men preferred their partners to self-administer treatments at home to avoid the costs and delays associated with facility-based care. Given a choice between 5-FU and Artesunate topical therapies, based on dosing frequency and other factors, a majority favored 5-FU for its biweekly application despite a required abstinence period. Focus group discussions revealed a preference for 5-FU’s perceived easier schedule and perceived higher effectiveness given its longer duration. However, some preferred Artesunate due to its daily dosing regimen and the absence of a mandatory abstinence period after use, which they believed could lead to better adherence.

Our study, to our knowledge, is the first to evaluate men’s perceived support of their female partners’ use of topical therapies for cervical precancer treatment in a sub-Saharan African country. The study’s findings of strong support from men for the use of topical therapies, including their willingness to follow recommendations for abstinence and condom use linked with these treatments, are promising for the potential use of these therapies in low- and middle-income countries (LMICs). Men’s support of the abstinence and condom use restrictions associated with these therapies are specifically crucial in sub-Saharan Africa and other LMICs, where some women may have reduced agency to negotiate these matters.^37^ Because of the pro-inflammatory nature of some intravaginal therapies, including 5FU, which can use cause transient erythema and edema of the vaginal mucosa, abstinence for two or three days following each application is usually recommended.^9,14,15^ In a few cases, superficial, self-limited erosions of the vaginal or epithelial mucosa have been observed.^9^ Inability to adhere to abstinence recommendations while using these therapies may be associated with worse side effects, potentially increasing a woman’s risk of contracting sexually transmitted infections, including HIV, and may expose the male partner to the agent in case of barrierless intercourse.^38^ Similarly, as some topical agents like 5FU are teratogenic when used systemically, women must use contraception during their use to avoid pregnancy. In the cervical precancer treatment literature, studies have found that while guidelines recommend abstinence for 4-6 weeks after ablation or excisional procedures,^32^ some women in Malawi,^39^ Kenya,^40^ and Peru ^41^ reported inability to maintain abstinence for the recommended period due to lack of male partner support.^38^ Further studies, including pilot trials of self-administered topical therapies in LMICs, can investigate men’s actual support of adherence recommendation, in contrast to the perceived support reported in our study. The shorter abstinence duration associated with topical therapies compared to conventional surgical precancer treatments may be an advantage for their use in these settings.

In both the qualitative and quantitative findings, study participants strongly supported the option of self-administration of topical therapies compared to provider-administration, specifically citing access limitations related to distance, cost, and long wait times associated with facility-based treatment. These challenges are well described in the literature.^4,5,42,43^ Significant loss-to-follow-up rates have been reported, as high as 40-50%, when women screened in rural facilities must visit central referral facilities to access treatment.^4,43^ In Malawi, between 2011-2015, of the 836 women requiring excisional treatment referred to central facilities, only 266 (31.8%) received treatment due to inadequate numbers of trained providers or lack of equipment.^5^ Self-administered topical therapies, whose efficacy has been demonstrated in studies in high-income settings,^10,13–15^ may be a feasible, innovative, and cost-effective way to bridge these precancer treatment gaps in LMICs.

In our study, participants expressed diverse opinions on which of two topical therapy options, differing in usage duration and requirements for contraception and abstinence, would be preferable to themselves or their female partners. In the surveys, more participants preferred 5FU, which was presented as used once every two weeks, to Artesunate, which is used daily, even though 5FU required abstinence for two days after use. In the FGDs, some men perceived this to be easier to remember compared to daily use. In contrast, in the FGDs, those who preferred Artesunate often cited its lack of abstinence restrictions, as well as a perception that daily dosing may be easier for women. Although these varied opinions come exclusively from men, they likely mirror diverse societal preferences for such products, underscoring the importance of providing options that cater to different contexts.

There are several limitations to our study. Our survey sample size of 214 participants, conveniently sampled over several months at outpatient clinics in western Kenya, may not be representative of men in other regions of Kenya and may not be generalizable to other settings. Participants were samples primarily from HIV clinics, explaining the high prevalence of self-reported HIV infection – 73 percent -in our study sample. It is possible that HIV-positive men, by virtue of having more interaction with the healthcare system, are more open to medical interventions like self-administered therapies compared to HIV-negative men. Similarly, as our survey responses are self-reported, despite the use of well-trained research assistants who attempted to normalize all participant responses, our high perceived acceptability may be affected by response bias. It is reassuring that the qualitative data largely support the quantitative findings, as most FGD participants affirmed their perceived support of their female partners using topical therapies, explaining how it topical therapies can address challenges associated with current treatments. Another limitation of our study is the non-comprehensive options of topical therapies presented to participants, where only two potential therapies were discussed, and reasons for participants’ preferences were not deeply probed. Future studies can employ the use of discrete choice experiments^44^ to gain further insights into different attributes of topical therapies that are valued by men in this setting.

In conclusion, we find high rates of self-reported acceptance of topical therapies for cervical precancer treatment among men of varied ages and socioeconomic status in Kenya. The vast majority of men, who were relatively well-informed about HPV and cervical cancer, expressed willingness to support their female partners in using a topical therapy to treat cervical precancer if it were accessible and recommended to them. Additionally, our quantitative and qualitative findings indicate a preference for self-administration at home over provider-administration in a health facility. This preference is mainly due to its potential to overcome obstacles women face in accessing facility-based treatments in resource-limited settings. Study participants also showed varied preferences regarding the attributes of topical therapies, including the frequency and duration of use, as well as the need for abstinence. Our findings offer important lessons for ongoing (Clinicaltrial.gov identifier NCT05413811, NCT05362955, NCT06165614) and future trials investigating the feasibility of repurposing topical therapies for cervical precancer treatment in LMIC settings.

## Data Availability

All data produced in the present study are available upon reasonable request to the authors

## List of Abbreviations

LMIC: low- and middle-income countries
5FU: 5-Fluorouracil
HPV: human papillomavirus
HIV: human immunodeficiency virus
WHO: World Health Organization
FGD: focus group discussion

## Financial declaration

This research was supported by the Eunice Kennedy Shriver National Institute of Child Health & Human Development of the National Institutes of Health under Award Number K12HD103085 and the Victoria’s Secret Global Fund for Women’s Cancers Career Development Award, in Partnership with Pelotonia Foundation and the American Association of Cancer Research (AACR). The content is solely the responsibility of the authors and does not necessarily represent the official views of the National Institutes of Health. The study funders had no role in the research.

## Authors Contributions

CM and LR conceptualized and designed the research and led manuscript writing, EA and MR supported implementation of the study, GE, YZ, KA performed the analysis and aided in data interpretation. All authors discussed the results and commented on the manuscript.

## Acknowledgements

The study team acknowledges all study participants and the support of the Kisumu County Ministry of Health and Lumumba Sub-County Hospital leadership.

---

**Supplemental Table 1.** Self-reported knowledge of HPV, cervical cancer, perceptions of risk, and attitudes toward female partner cervical cancer screening and treatment among partnered men in Kisumu County, Kenya, n=214

| Question | Yes/True (%) | No/False (%) | Unsure (%) |
| --- | --- | --- | --- |
| Heard of cervical cancer previously | 210 (98) | 4 (2) | 0 (0) |
| Heard that women can screen for cervical cancer | 209 (98) | 4 (2) | 1 (0.5) |
| Know anyone with cervical precancer or cancer | 72 (34) | 141 (66) | 1 (0.5) |
| Heard of human papillomavirus before today | 81 (38) | 132 (62) | 1 (0.5) |
| HPV can be transmitted via sexual intercourse | 69 (85) | 3 (4) | 9 (11) |
| Both men and women can be infected with HPV | 51 (63) | 22 (27) | 8 (10) |
| HPV infection is always symptomatic | 36 (44) | 36 (44) | 9 (11) |
| HPV can cause cervical cancer | 78 (96) | 0 (0) | 3 (4) |
| Having HIV increases a woman's risk of getting HPV or cervical cancer | 73 (90) | 3 (4) | 5 (6) |
| Smoking cigarettes increases risk of getting HPV or cervical cancer (n=158) | 97 (61) | 19 (12) | 42 (27) |
| Cervical cancer is preventable via vaccination or screening | 163 (76) | 22 (10) | 29 (14) |
| Only women with symptoms should get screened for cervical cancer | 27 (13) | 185 (86) | 2 (1) |
| Cervical cancer can be treated or cured if diagnosed early | 211 (99) | 2 (1) | 1 (0.5) |
| Partner may be at risk for cervical cancer | 139 (65) | 50 (23) | 25 (12) |
| Partner has undergone cervical cancer screening | 179 (84) | 26 (12) | 9 (4) |
| Would support cervical cancer screening in partner | 212 (99) | 1 (0.5) | 1 (0.5) |
| Would support treatment of cervical precancer or cancer in partner | 211 (99) | 2 (1) | 1 (0.5) |
| Diagnosis of cervical precancer or cancer would negatively affect relationship | 5 (2) | 206 (96) | 3 (1) |

